# Application of Impulse Oscillometry Combined with Bronchodilator Testingfor Asthma Management: Children’s Hospital Experience in Taiwan

**DOI:** 10.64898/2026.01.30.26345207

**Authors:** I-Han Cheng, Hsiang-En Lin, Wen-Jue Soong, Ti-Chien Lu, Su-Boon Yong, Chien-Heng Lin

**Author notes:** Corresponding: Su-Boon Yong MD, PhD., Department of Allergy and Immunology, China Medical University, Children’s Hospital, Taichung, Taiwan, Research Center for Allergy, Immunology, and Microbiome (A.I.M.), China Medical University, Children’s Hospital, Taichung, Taiwan, Chien-Heng Lin, MD, Division of Pediatric Pulmonology and Critical Medicine, China Medical University Children’s Hospital, No. 2, Yuh-Der Road, Taichung 404, Taiwan. **Ethics approval** This study was approved by the Research Ethics Committee I of China Medical University Hospital, Taichung, Taiwan (Approval No. CMUH109-REC1-024). The protocol was approved by **Martin M-T Fu, MD, DMSc**, Chairman, Research Ethics Committee I, China Medical University Hospital.

## Abstract

**Background:** Impulse oscillometry is a noninvasive pulmonary function test performed during quiet breathing and requires minimal patient cooperation. It is useful for detecting small airway disease and provides increased sensitivity for diagnosing asthma in younger children who may have difficulty completing standard spirometry. Bronchodilator testing, a standard assessment of airflow obstruction reversibility, is recommended in patients with suspected asthma who present obstructive airflow patterns.

**Objective:** To evaluate impulse oscillometry parameters before and after bronchodilator administration across different age groups and to examine the relationship between age and airway resistance in patients with clinician-diagnosed asthma.

**Methods:** This retrospective study included patients with clinician-diagnosed asthma who demonstrated obstructive airflow patterns and a positive bronchodilator response. Participants were grouped by age: younger than 6 years, 6 to 20 years, and older than 20 years. Key impulse oscillometry parameters—airway resistance at 5 Hz, airway resistance at 20 Hz, the difference between these values, and resonance frequency—were collected and compared across groups. A positive bronchodilator response was defined as a reduction in airway resistance of more than 30% in individuals younger than 18 years and more than 40% in adults.

**Results:** A total of 225 patients (123 males and 102 females) were included, with a median age of 6 years. At baseline, the median airway resistance at 5 Hz was 175.34% of the reference value (95% CI, 171.66–178.62), and airway resistance at 20 Hz was 121.68% (95% CI, 118.73–127.12). The median difference between these values was 52.32% (95% CI, 49.89–57.14), and the median resonance frequency was 5.11 Hz (95% CI, 4.62–5.35). After bronchodilator administration, airway resistance at 5 Hz decreased to 123.56% (95% CI, 119.07–126.77), corresponding to a median reduction of 52.8% (95% CI, 49.48–56.08; P < 0.0001). Age demonstrated a moderate positive correlation with airway resistance at 20 Hz (r = 0.51, P < 0.001).

**Conclusions:** Proximal airway resistance increases with age among patients with asthma, suggesting age-related differences in airway inflammation. Impulse oscillometry combined with bronchodilator assessment provides a practical approach for evaluating airflow reversibility and enhances diagnostic accuracy in suspected asthma.

## Introduction

Asthma is a chronic respiratory condition characterized by inflammation and hyperresponsiveness of the airways, significantly impacting the lives of millions of children worldwide(1). Despite advancements in treatment(2), the accurate diagnosis and effective management of asthma in pediatric patients remain challenging(3). Traditional pulmonary function tests (PFTs), such as spirometry, require substantial patient cooperation, which can be difficult to obtain from young children(4-6). This limitation often results in underdiagnosis or misdiagnosis, hindering proper asthma management(4-6).

The need for more reliable and patient-friendly diagnostic tools is evident, particularly for pediatric populations who struggle with standard PFTs(4). Impulse oscillometry (IOS) has emerged as a promising alternative, offering non-invasive assessment of respiratory function that requires minimal patient effort(6-9). However, there is a gap in the literature regarding the effectiveness of IOS combined with bronchodilator tests (BDT) in improving asthma diagnosis and management among children(6, 9-11). Addressing this gap is crucial for optimizing asthma care and improving long-term health outcomes for pediatric patients.

The primary objective of this study is to evaluate the effectiveness of IOS combined with BDT in managing asthma among children at China Medical University Children’s Hospital. Specifically, the study aims to: 1) assess the reduction in airway resistance post-bronchodilator administration, 2) investigate correlations between respiratory parameters and patient age, and 3) determine the practicality and sensitivity of IOS in detecting small airway disease in young children.

Previous studies have demonstrated the potential of IOS in diagnosing respiratory conditions and monitoring treatment efficacy(6, 9, 12, 13). IOS has shown greater sensitivity in detecting small airway disease and has been particularly useful in populations that struggle with standard PFTs(14, 15). Despite these findings, limited research has focused on its application in routine clinical practice for pediatric asthma management, highlighting the need for further investigation.

This study posits that the incorporation of IOS combined with BDT into the diagnostic process for pediatric asthma provides a more accurate and non-invasive alternative to traditional PFTs, leading to improved asthma management and patient outcomes.

## Material and Methods

### Study subjects

The hospital’s institutional review board (IRB) concurred that this retrospective study was a continuous quality improvement initiative to improve patient care and did not require informed consent. This study was approved by the IRB of our hospital (**CMUH109-REC1-024**).

Patients with asthma received IOS between August 1, 2019 and May 31, 2020 were retrospectively enrolled. All included patients demonstrated obstructive airflow on spirometry and a spirometry-defined positive bronchodilator test (BDT). A positive BDT was defined according to ATS/ERS criteria as an increase of ≥12% and ≥200 mL in FEV □ following administration of 400 μg salbutamol. Asthma diagnosis followed the Global Initiative for Asthma (GINA) guidelines. IOS parameters were collected independently and were not used to determine BDT positivity. Asthma was diagnosed according to GINA guidelines. The exclusion criteria were as follows: patients had medications of asthma controller recent 2 days, patients who had congenital heart disease, or had co-existing other pulmonary anomality, such as bronchopulmonary dysplasia or pulmonary neoplasm.

Patients were grouped by age less than 6-year-old, 6 to 20 year-old, and above 20 year-old. Data of R5, R20, R5-R20, resonance frequency were collected, analyzed and compared between these 3 groups. These cutoffs were selected because IOS performance and airway physiology differ substantially between preschool children (<6 years), school-aged children and adolescents (6–20 years), and adults (>20 years), consistent with previous IOS and lung development literature. Preschool children often cannot reliably perform spirometry, while older children and adults generally can, making these groups clinically distinct for interpretation. Reduction of airflow resistance by more than 30% in children younger than 18 year-old or 40% in adults after BDT is defined positive findings in our study. These thresholds were consistently applied to classify BDT positivity for all subjects, and all subsequent analyses of bronchodilator responsiveness were based on these predefined criteria.

Impulse oscillometry (IOS) was performed using the Jaeger MasterScreen system (Erich Jaeger, Höchberg, Germany) in accordance with the ATS/ERS Technical Standards for Respiratory Oscillometry (16). All patients underwent IOS prior to spirometry as part of routine clinical testing. During the procedure, patients were seated, supported their cheeks and the floor of the mouth with their hands to minimise upper airway shunting, and were instructed to breathe normally while the loudspeaker generated pressure impulses for approximately 20 seconds. A disposable mouthpiece and nose clip were used for all measurements.For each participant, the bronchodilator response was quantified using paired change scores (Δ), calculated as the difference between post-BDT and pre-BDT IOS values (Δ = Post – Pre). This ensured that each patient served as their own control. All comparisons of pre- and post-BDT parameters were therefore performed using paired statistical analyses. R5%, R20%, X5%, and other IOS parameters expressed as percentages represent “percent of predicted” values calculated automatically by the MasterScreen system based on the device’s built-in reference equations(17). Raw resistance values (in hPa·s/L) were also recorded by the system.

### ANALYSIS

Continuous variables were expressed as median ± IQR (interquartile range) and were analyzed using the Wilcoxon signed-rank test. The relationship between age and airway resistance was assessed using the Pearson correlation coefficient. The SPSS package, version 12 (SPSS, Inc., Chicago, IL, USA) was used for the analyses, and p-values less than 0.05 were considered statistically significant for rejecting the null hypothesis.

#### Ethical Considerations

This retrospective study was conducted at **China Medical University Children’s Hospital, Taichung, Taiwan**. The hospital’s institutional review board (IRB) concurred that this study was a continuous quality improvement initiative aimed at improving patient care and therefore did not require informed consent. The study was approved by the IRB of China Medical University Hospital (CMUH109-REC1-024).

## Results

A total of 225 patients were enrolled in the study between August 1, 2019, and May 31, 2020. The mean age was 6 years (±2.3), with an age range of 2 to 23 years. Of these, 126 (54.67%) were male, and 102 (45.33%) were female.

Table 1 shows that both Pre-R5% and Post-R5% values were obtained for all 225 patients. The lowest recorded Pre-R5% was 1.58%, while the lowest Post-R5% was 1.07%. The highest Pre-R5% was 458.09%, compared to 291.09% for Post-R5%. The median Pre-R5% was 175.34%, while the median Post-R5% was 123.56%. The 95% confidence intervals were 171.66% to 178.62% for Pre-R5% and 119.07% to 126.77% for Post-R5%. The interquartile range was 164.43% to 197.28% for Pre-R5% and 107.59% to 141.92% for Post-R5%.

**Table 1.**
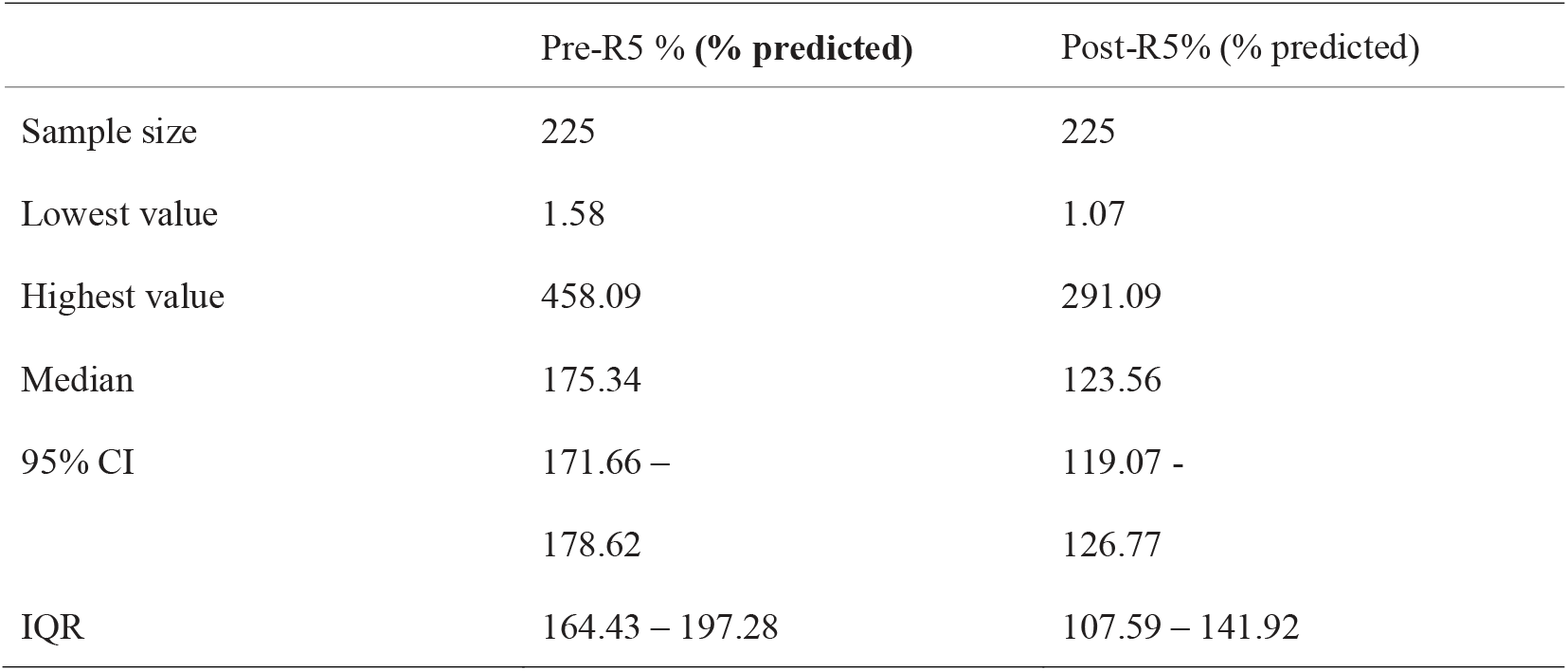
Figure 1. Comparison of pre-BD and post-BD R5 values expressed as percent of predicted (% predicted). Error bars indicate 95% confidence intervals.

This table illustrates a significant reduction in airway resistance following bronchodilator therapy, as indicated by the lower post-treatment values.

Figure 1 displays the Pre-R5% and Post-R5% airway resistance values. The taller bar on the left represents Pre-R5%, indicating higher resistance before treatment, while the shorter bar on the right represents the lower Post-R5% values after treatment. Both bars include error bars representing 95% confidence intervals, with red dots marking the mean values. The Pre-R5% bar shows a value close to 175%, and the Post-R5% bar is near 125%, confirming a marked reduction in airway resistance after bronchodilator administration.

**Figure 1.**
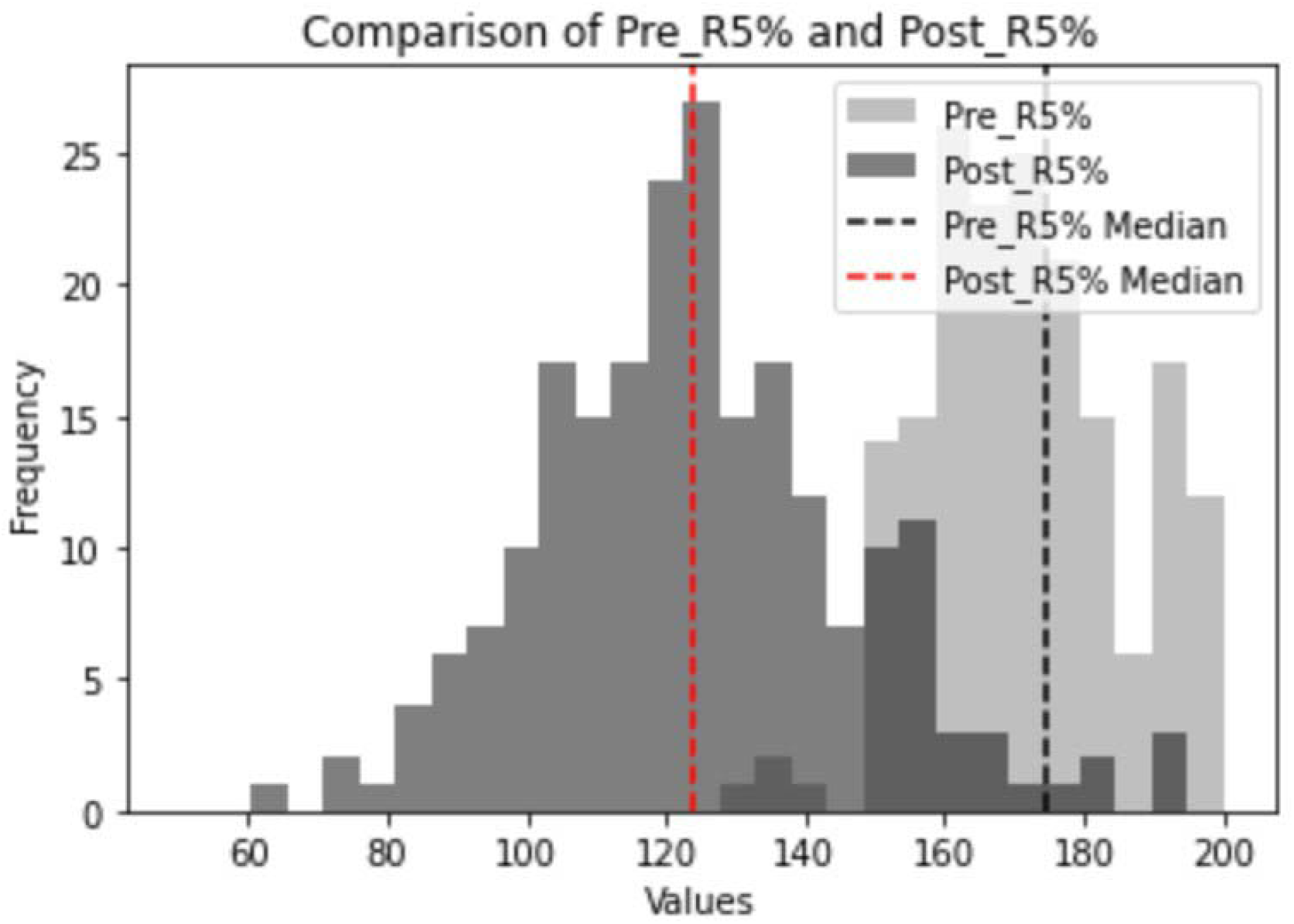
The bar chart compares pre-treatment (Pre-R5%) and post-treatment (Post-R5%) airway resistance percentages. Here are the key observations from the chart:

**Figure 2.**
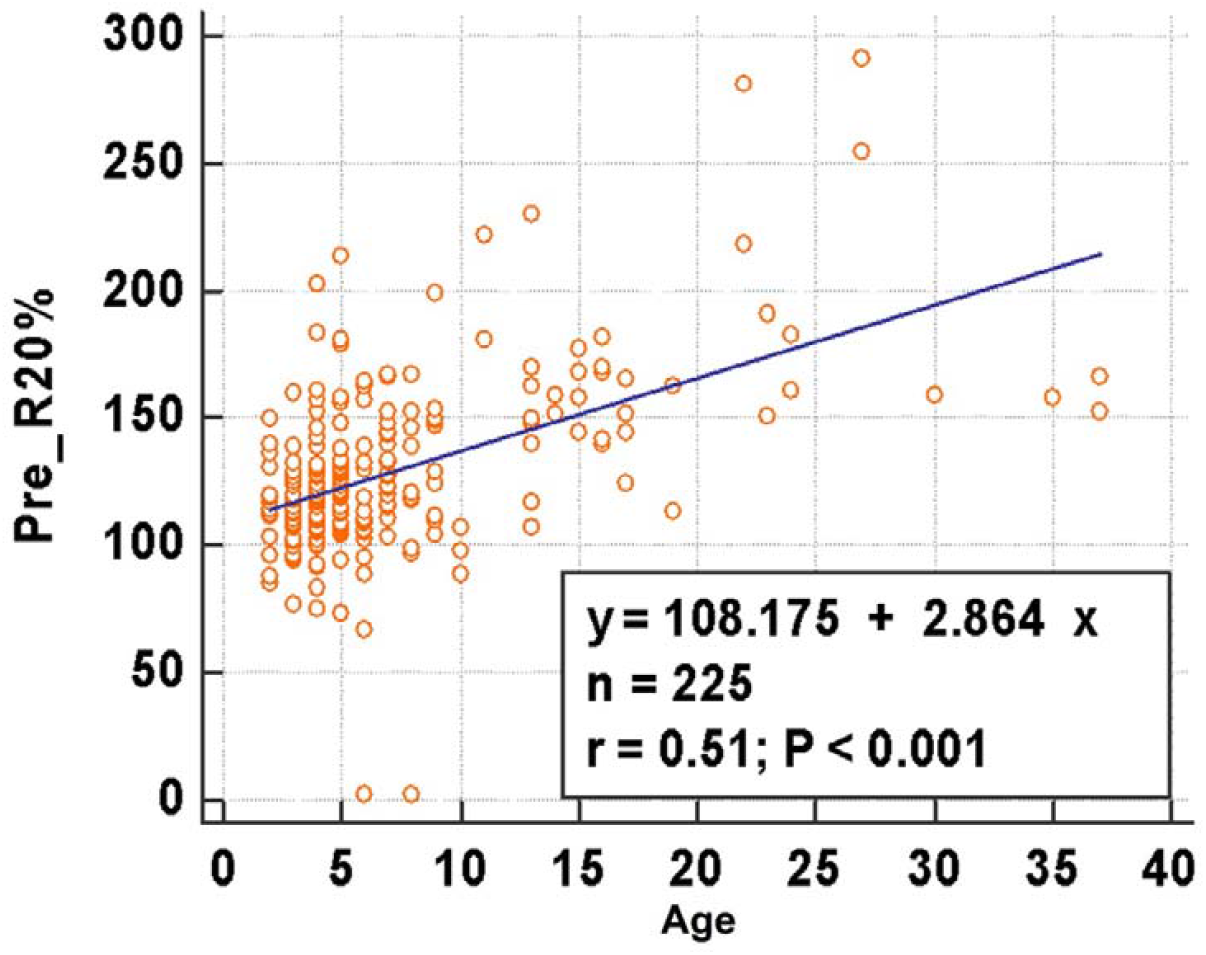
There is moderate correlation between age and R20 values (r=0.51, P < 0.001).

Applying our predefined cutoff criteria, the median reduction in R5 (–52.8%) exceeded the 30% threshold for a positive BDT response in children, indicating that the majority of participants demonstrated a clinically meaningful bronchodilator response. Although adults comprised a small proportion of the sample, their reductions also exceeded the 40% adult threshold, confirming consistent application of the positivity definitions across age groups.

Correlations between age and IOS parameters are shown in Figure.2 but weak correlation in pre-BD R5 (r=0.16, P=0.014), post-BD R5(r=0.24, *p* <0.001), R5-R20 (r=0.28, *p* < 0.001), Resonance frequency (r=0.30, P<0.001) but no correlation in X5(r=0.09, *p*=0.319) and AX5(r=0.05, *p* =0.423). Age showed a weak but statistically significant correlation with both pre-BD R5 (r = 0.16, p = 0.014) and post-BD R5 (r = 0.24, p < 0.001), consistent with previous literature.

These age-related correlations were exploratory and were not associated with the magnitude of bronchodilator responsiveness. Other IOS parameters (X5, AX, Fres, R5–R20, R20) did not demonstrate clinically meaningful ability to define bronchodilator response beyond the changes observed in R5.

## Discussion

This study aimed to evaluate the effectiveness of IOS combined with BDT in the management of asthma among children. By analyzing data from 225 patients at China Medical University Children’s Hospital, we found that IOS is a valuable tool for diagnosing and monitoring asthma, especially in children who may not be able to perform standard pulmonary function tests (PFTs). The significant reduction in airway resistance observed post-BDT values confirm the utility of IOS in assessing reversible airflow obstruction in young children with asthma.

Previous studies have suggested that asthma can be reliably diagnosed with the following IOS bronchodilator response values: a 20%-40% deceased in R5 and a 15% to 30% decreased in R10 (16). Our findings underscore the practicality and sensitivity of IOS in assessing bronchodilator responsiveness in paediatric asthma.. The ability of IOS to measure respiratory impedance during tidal breathing with minimal patient cooperation makes it an ideal method for pediatric patients. The significant reduction in R5 values post-BDT (from a median of 175.34% to 123.56%) highlights the effectiveness of BDT and reinforces the importance of IOS in asthma management protocols.

The study revealed a moderate correlation between age and R20 values, indicating that proximal airway resistance increases with age. While this may reflect age-related differences in airway calibre and developmental physiology, these changes did not influence bronchodilator responsiveness. In other words, although age correlated with several baseline IOS parameters, the magnitude of BDT response remained age-independent, indicating that the observed bronchodilator effects were not driven by age-related physiological variation.

Although age correlated with several baseline IOS parameters, our study did not measure airway inflammation or structural changes, and therefore no mechanistic conclusions can be drawn about age-related inflammatory differences between children and adults. From a physiological perspective, this pattern is consistent with age-related changes in airway calibre and respiratory mechanics. As children grow into adolescence, increases in lung volume and airway dimensions, as well as maturation of the chest wall, can alter the relative contribution of the central airways to overall respiratory impedance. In younger children, higher R20 values may partly reflect smaller airway calibre and developmental anatomy, whereas in older children and adults, persistently elevated R20 despite growth could be interpreted as a more clinically relevant marker of proximal airway involvement. These age-related considerations support the need for age-aware interpretation of IOS indices in routine practice.

By analysing individual paired changes rather than comparing pooled pre- and post-BDT values, we ensured that bronchodilator responsiveness reflected true within-patient effects. The substantial negative ΔR5% confirmed that airway resistance consistently decreased after BDT administration. Although age correlated with several baseline IOS parameters, these correlations did not influence ΔR5% or the magnitude of bronchodilator response. Furthermore, IOS indices other than R5 showed smaller and more variable Δ values, indicating that R5 remains the most sensitive IOS marker for detecting bronchodilator responsiveness in this paediatric cohort.Previous studies have highlighted the usefulness of IOS in correlating respiratory measurements with age, sex, and height.(33-35) Consistent with previous literature, our study also showed weak but statistically significant correlations between age and R5 values in both pre- and post-bronchodilator measurements.. These correlations further validate the use of IOS as a reliable diagnostic tool for assessing asthma severity and treatment response in children.

Despite its advantages, IOS has limitations. It does not provide information on lung volumes or oxygen diffusion capacity, which are critical in a comprehensive asthma assessment(36, 37). Additionally, the lack of universally accepted reference values for IOS parameters across different patient populations may limit its generalizability(5, 16). Future research should focus on establishing standardized reference values and exploring the integration of IOS with other diagnostic modalities to enhance its diagnostic accuracy.

Moreover, our study was retrospective, which may introduce selection bias and was lack of control group. Prospective studies with larger sample sizes and diverse patient demographics are needed to validate our findings and further explore the clinical utility of IOS combined with BDT in asthma management.

The findings of this study have several implications for clinical practice. First, the consistent reduction in R5 following BDT supports the routine use of IOS as an accessible and child-friendly tool for assessing reversible airflow obstruction in paediatric asthma, particularly in settings where spirometry is challenging or not feasible. Second, the moderate age-related increase in R20 highlights the need for age-aware interpretation of IOS parameters when evaluating airway resistance in clinical settings. For future research, prospective longitudinal studies are needed to establish age-specific IOS reference ranges, validate optimal bronchodilator response cutoffs, and determine whether IOS-based markers can predict long-term asthma control or exacerbation risk. Integrating IOS with biomarkers or imaging may further enhance its diagnostic and prognostic capabilities in paediatric asthma.

Our study supports the integration of impulse oscillometry (IOS) with bronchodilator testing (BDT) into routine diagnostic protocols for asthma as a feasible and effective method for assessing reversible airflow obstruction, especially in young children who may struggle with standard pulmonary function tests. The findings also demonstrate that proximal airway resistance increases with age in asthmatic patients, highlighting the importance of age-aware interpretation of IOS parameters across childhood, adolescence and adulthood. The bronchodilator response of R5, reflecting a significant reduction in airway resistance after BDT, emerges as the most reliable IOS parameter for confirming asthma diagnosis in clinical practice. Moreover, older patients exhibit higher proximal airway resistance, as indicated by elevated R20 values. Given these findings, IOS should be considered an essential tool in asthma diagnosis and management, particularly for young children who face challenges with conventional pulmonary function tests. By enhancing diagnostic precision and monitoring, IOS may contribute to improved outcomes for pediatric asthma patients.

## Data Availability

All data produced in the present study are available upon reasonable request to the authors

## Assistance with the study

None.

## Presentation

None.

